# A study to assess the impact of cobas Liat point-of-care PCR assays (SARS-CoV-2 and Influenza A/B) on patient clinical management in the emergency department of the University of California at Davis Medical Center

**DOI:** 10.1101/2022.08.08.22278536

**Authors:** Larissa May, Elissa M. Robbins, Jesse A. Canchola, Kamal Chugh, Nam K. Tran

## Abstract

**Background:** Rapid detection of SARS-CoV-2 is crucial for reduction of transmission and clinical decision-making. The cobas^®^ SARS-CoV-2 & Influenza A/B nucleic acid test for use on the cobas Liat^®^ System is a rapid (20 minutes) point-of-care (POC) polymerase chain reaction (PCR) method.

**Methods:** This unblinded, pre-post study enrolled consecutive patients with symptoms/signs consistent with SARS-CoV-2 infection presenting to the University of California, Davis emergency department (ED). Outcomes following implementation of the cobas Liat SARS-CoV-2 & Influenza A/B test (intervention period: December 2020–May 2021) were compared with previous standard-of-care using centralized laboratory PCR methods (control period: April 2020–October 2020).

**Results:** Electronic health records of 8879 symptomatic patients were analyzed, comprising 4339 and 4540 patient visits and 538 and 638 positive SARS-CoV-2 PCR test results in the control and intervention periods, respectively. Compared with the control period, turnaround time (TAT) was shorter in the intervention period (median 0.98 vs 12.3 hours; p<0.0001). ED length of stay (LOS) was generally longer in the intervention period compared with the control period, but for those SARS-CoV-2-negative who were admitted, ED LOS was shorter (median 12.53 vs 17.93 hours; p<0.0001). Overall, the rate of anti-infective prescribing was also lower in the intervention period than in the control period (antibiotics only: 38.11% vs 44.55%; p<0.0001 and antivirals only: 3.13% vs 0.94%; p<0.0001).

**Conclusion:** This real-world study confirms faster TAT with a POC PCR method in an emergency care setting and highlights the importance of rapid SARS-CoV-2 detection to aid patient management and inform treatment decisions.

**Clinical Relevance:** This study reports data collected from a quasi-experimental pre-post study using the electronic health records of patients presenting to the emergency department (ED) of the University of California at Davis Medical Center with symptoms or signs consistent with SARS-CoV-2 infection during their ED visit. The primary objective of this study was to determine if implementation of the point-of-care (POC) cobas^®^ Liat^®^ SARS-CoV-2 & Influenza A/B test for use on the cobas Liat System reduced the diagnostic turnaround time and/or length of stay for ED patients with suspected SARS-CoV-2 infection compared with the previous standards of care (batch-wise diagnostic testing using the cobas 6800 System and on-demand urgent testing on the GenMark Dx^®^ ePlex^®^ system in a centralized clinical laboratory). Ultimately, these data help to inform how implementation of POC molecular testing methods impact patient management.

## Introduction

As of May 26, 2022, the coronavirus disease (COVID-19) pandemic has resulted in an estimated 83,408,645 cases of COVID-19 in the United States (US), and 1,000,254 deaths (1). Even as vaccination levels increase in the general population, testing of symptomatic individuals is vital for controlling the spread of SARS-CoV-2 (severe acute respiratory syndrome coronavirus 2; the causative agent of COVID-19), as vaccination alone does not prevent infection and transmission (2, 3). The choice of diagnostic test (e.g., nucleic acid amplification test [NAAT] or antigen detection and point-of-care [POC] or centralized testing) is setting dependent (4). Resources (e.g., availability of trained staff), turnaround time (TAT), testing capacity, assay performance (sensitivity and specificity), and patient characteristics, such as days since symptom onset, all need to be considered (4).

Vaccination has resulted in the relaxation of public health mitigation measures in many geographical regions (5-7). The increase in respiratory viruses that had hitherto been reduced due to physical distancing measures (8, 9) makes it important for a clinician to determine whether COVID-19-associated symptoms are attributable to SARS-CoV-2 infection, another respiratory infection (4), or a non-infectious etiology. The SARS-CoV-2 Omicron (B.1.1.529) variant, which has a shorter incubation period than the Alpha (B.1.1.7) variant (10) and potentially higher transmissibility than the ancestral viruses (11), increases the need for rapid diagnosis. As the pandemic has evolved, monoclonal antibody therapies have become available that are more effective if administered in the early stages of symptomatic infection (12), meaning that rapid diagnosis also has important implications for treatment.

In the emergency department (ED), test results are often not immediately available (13); typical TATs for high-throughput NAATs can range from ∼1.5 hours to 8 hours (14), with approximately 97% of polymerase chain reaction (PCR) results available within 23 hours from arrival in the ED (13). Symptomatic patients should be isolated in an examination room or designated area, in the absence of a laboratory-confirmed diagnosis, with consequences for allocations of hospital resources, including the need for isolation bays and personal protective equipment (15, 16). Given the implications for infection prevention, and the variability in isolation capacity (16), rapid results are helpful for the effective triage of ED patients with suspected SARS-CoV-2, both to reduce transmission and to identify those that may benefit from hospital admission or other therapeutic intervention (4). Further, performing on-demand POC testing in the ED while using centralized high-throughput systems for less urgent testing needs could increase overall testing capacity.

At the onset of this study, the University of California (UC) at Davis Medical Center performed diagnostic testing in a centralized clinical laboratory. The cobas^®^ Liat^®^ SARS-CoV-2 & Influenza A/B nucleic acid test for use on the cobas Liat System (cobas Liat SARS-CoV-2 & Influenza A/B test) is intended for the simultaneous rapid *in vitro* detection and differentiation of SARS-CoV-2 and influenza A and B virus nucleic acids in clinical specimens from individuals suspected of respiratory viral infection (17). The assay provides results from a single test to rule-in or rule-out influenza and COVID-19 simultaneously, within approximately 20 minutes (17).

The primary objective of this study was to determine if implementation of the cobas Liat SARS-CoV-2 & Influenza A/B test reduced the diagnostic assay TAT and/or length of stay (LOS) for ED patients with suspected SARS-CoV-2 infection compared with the previous standards of care. The secondary objectives were to determine the impact of POC testing on ED disposition and the use of anti-infectives in patient management.

## Materials and Methods

### Study design

This was a retrospective, unblinded, quasi-experimental pre-post study The study comprised two periods: a control period (April 01, 2020 to October 31, 2020) and an intervention period (December 01, 2020 to May 31, 2021).

### Participants

Individuals presenting at the UC Davis ED were included in the study consecutively if they had symptoms or signs consistent with COVID-19 at their ED visit (**Supplementary Table S1**). A real-time reverse transcriptase PCR (RT-PCR) SARS-CoV-2 test must also have been ordered by the ED physician during the patient’s ED visit, in either study period. For each patient included in the study, the information of a complete event, defined as all diagnostics and prescriptions ordered within 30 days of the initial ED visit, was collected using electronic health records.

### Sample collection, storage and testing

Samples were collected according to manufacturer instructions and analyzed as soon as possible, not exceeding maximum storage times and temperatures dictated by the manufacturer’s instructions for use (IFU) (17-19). In the control period, centralized diagnostic testing was conducted using the cobas SARS-CoV-2 test on the cobas 6800 System (19) or on-demand urgent testing using the GenMark Dx^®^ ePlex^®^ SARS-CoV-2 Test (18). The cobas 6800 System can perform more than 1000 tests per day, with first results reported in approximately 3 hours of loading (20, 21). In contrast, the assay run time for the ePlex SARS-CoV-2 Test is approximately 90 minutes (22) allowing for up to 288 samples per day (23) depending on instrument configuration and lab staffing. In the intervention period, the cobas Liat SARS-CoV-2 & Influenza A/B test was used for detection of the target sequences (17).

### Analysis

For the purpose of this analysis, data from the two centralized testing methods were pooled to allow the comparison between POC PCR and centralized PCR methods. Data collected during November 2020 were excluded from the analysis as this was considered a transition period, allowing a phasing in of the POC assay. Samples yielding indeterminate/invalid results were handled per the assay IFUs (17-19). More information on test methods and analysis is available in the **Supplementary Materials**.

### Statistical analysis

No formal sample size calculations were conducted for this study. All data analyses were performed using SAS/STAT^®^ v9.4 software (24). Comparisons were made at the patient visit-level rather than continuously over the study duration. Patient data included in this study were summarized using descriptive statistics and compared between the control and intervention periods. The primary endpoints of TAT and LOS, and the secondary endpoints of ED disposition and anti-infective prescription rates, were also analyzed by SARS-CoV-2 test result. A post hoc analysis of LOS and anti-infective use was conducted to stratify data by ED disposition and by SARS-CoV-2 test result and ED disposition.

Statistical hypothesis testing was performed using the Chi-Square test for categorical data, the non-parametric Wilcoxon rank-sum test for continuous-level non-normally distributed data and the Student’s t-test for normally distributed data. The rate of type I errors was controlled for using the false discovery rate method when conducting multiple comparisons. Indeterminate/invalid results were combined and included in the relevant tables but were not included in statistical hypothesis testing due to small sample sizes.

### Ethics statement

This study was conducted in compliance with the International Conference on Harmonisation Good Clinical Practice Guidelines, and applicable US Food and Drug Administration regulations. The study protocol and de-identification procedure for patient data were approved by the UC Davis Institutional Review Board Administration.

### Data availability

The data used to support the findings of this study are included within the article and supplementary materials.

## Results

### Patient demographics and characteristics

A total of 8879 medical records were included in the study (**Supplementary** Error! Reference source not found.). Patient demographics were similar (all comparisons, p>0.05) in the control and intervention cohorts (**Table 1**) and were therefore not controlled for in subsequent analyses. Across both periods, mean patient age was 47 years (median 51 years), the majority were White (42.03%), and 48.64% were female. Almost 40% of patients had a body mass index below the cut-off for obesity (<30.0 kg/m^2^) (25).

**Table 1:** Patient demographics

|  | Control period<br>(Centralized PCR) | Intervention period<br>(POC PCR) | p-value |
| --- | --- | --- | --- |
| **Total patient visits, n** | **4339** | **4540** |  |
| **Age (years), n (%)** |  |  | 0.63<sup>a</sup> |
| ≤30 | 1152 (26.55%) | 1259 (27.73%) |  |
| 31–40 | 492 (11.34%) | 505 (11.12%) |  |
| 41–50 | 481 (11.09%) | 485 (10.68%) |  |
| 51–60 | 691 (15.93%) | 685 (15.09%) |  |
| 61–70 | 723 (16.66%) | 727 (16.01%) |  |
| 71–80 | 488 (11.25%) | 526 (11.59%) |  |
| ≥81 | 312 (7.19%) | 353 (7.78%) |  |
| **Age (years)** |  |  | 0.18<sup>b</sup> |
| Mean (SD) | 47.47 (24.17) | 46.76 (25.34) |  |
| Median (IQR) | 51 (29, 66) | 51 (28, 67) |  |
| Range | (0, 90) | (0, 90) |  |
| **BMI<sup>c</sup> (kg/m<sup>2</sup>), n (%)** |  |  | 0.11<sup>a</sup> |
| Below 18.5 (underweight) | 236 (5.44%) | 270 (5.95%) |  |
| 18.5–24.9 (healthy) | 767 (17.68%) | 711 (15.66%) |  |
| 25.0–29.9 (pre-obesity) | 756 (17.42%) | 680 (14.98%) |  |
| 30–39.9 (obesity class I) | 726 (16.73%) | 736 (16.21%) |  |
| ≥40 (obesity class II) | 251 (5.78%) | 259 (5.70%) |  |
| Unknown<sup>e</sup> | 1579 (36.39%) | 1850 (40.75%) |  |
| **BMI<sup>c</sup> (kg/m<sup>2</sup>)** |  |  | 0.56<sup>b</sup> |
| Mean (SD) | 28.47 (8.22) | 28.60 (8.59) |  |
| Median (IQR) | 27.4 (22.7, 32.8) | 27.4 (22.4, 33.5) |  |
| Range | (14.0, 65.1) | (14.0, 65.5) |  |
| **Sex, n (%)** |  |  | 0.30<sup>a,d</sup> |
| Female | 2086 (48.08%) | 2233 (49.19%) |  |
| Male | 2251 (51.88%) | 2305 (50.77%) |  |
| Other | 2 (0.05%) | 2 (0.04%) |  |
| **Ethnicity, n (%)** |  |  | 0.24<sup>a,d</sup> |
| Hispanic/Latino | 969 (22.33%) | 972 (21.41%) |  |
| Not Hispanic/not Latino | 3318 (76.47%) | 3534 (77.84%) |  |
| Unknown<sup>e</sup> | 50 (1.15%) | 34 (0.75%) |  |
| Not reported | 2 (0.05%) | 0 (0.00%) |  |
| **Race, n (%)** |  |  | 0.12<sup>a, d</sup> |
| American Indian/Alaskan Native | 46 (1.06%) | 47 (1.04%) |  |
| Asian | 342 (7.88%) | 408 (8.99%) |  |
| Black/African-American | 836 (19.27%) | 936 (20.62%) |  |
| Native Hawaiian/Pacific Islander | 103 (2.37%) | 84 (1.85%) |  |
| White | 1843 (42.48%) | 1889 (41.61%) |  |
| Multiple/other | 1104 (25.44%) | 1133 (24.96%) |  |
| Not reported | 65 (1.50%) | 43 (0.95%) |  |

This table may include multiple visits per patient. All individuals with symptoms of a possible SARS-CoV-2 infection, and a PCR SARS-CoV-2 test ordered by a clinician during the control period (April 2020–October 2020) or the intervention period (December 2020– May 2021), were considered eligible for this study and are included in this summary table.
<sup>a</sup>Chi-square test
<sup>b</sup>Student's t-test
<sup>c</sup>N=58 observations were excluded from the BMI analysis due to data quality issues. BMI cut-off based on Centers for Disease Control and Prevention definitions (25)
<sup>d</sup>Chi-square test excludes 'Other', 'Unknown', or 'Not reported' categories
<sup>e</sup>Unknown category indicates subjects for whom the corresponding information is not available or reported as unknown
BMI, body mass index; IQR, interquartile range; PCR, polymerase chain reaction; POC, point-of-care; SARS-CoV-2, severe acute respiratory syndrome coronavirus 2; SD, standard deviation

The most commonly recorded SARS-CoV-2-related symptom was shortness of breath/difficulty breathing (21.83% control period, 18.57% intervention period) (**Supplementary Table S1**). Only one symptom was recorded for 54.51% of patients in control period and 47.97% in the intervention period), with 2 or ≥3 symptoms reported by <3% of patients each in either period (**Supplementary Table S2**).

More SARS-CoV-2 PCR tests were conducted in the intervention period than in the control period (n=4540 vs n=4339), and of those, more tested positive in the intervention period (14.05% vs 12.40%, respectively). Twelve (0.28%) tests were recorded as invalid/other in the control period versus none in the intervention period (**Supplementary Table S3**; p<0.02 overall for test outcome between periods).

### Impact of rapid POC testing on TAT

The TAT (defined here as time from electronic SARS-CoV-2 test order to result reporting) was shorter in the intervention period compared with the control period (median 0.98 vs 12.3 hours; p<0.0001) (**Table 2**). The lower end of the range for TAT in the control period (range 1.92, 74.38 hours) likely reflects samples analyzed with the ePlex SARS-CoV-2 Test, which has a shorter TAT than the cobas 6800. TAT was similarly reduced in the intervention period when analyzed by SARS-CoV-2 test result (**Table 2**) and when TAT was calculated as the time from sample collection to result reporting (**Supplementary Table S4**).

**Table 2:** Summary of SARS-CoV-2 test turnaround time (TAT), order to result

|  | **Control period** | **Intervention period** | **p-value<sup>a</sup>** |
| --- | --- | --- | --- |
|  | **(Centralized PCR)** | **(POC PCR)** |  |
| **Total patient visits, n** | **4339** | **4540** |  |
| **TAT<sup>b</sup> (hours)** |  |  | **<0.0001<sup>c</sup>** |
| Mean (SD) | 13.32 (6.77) | 1.29 (1.79) |  |
| Median (IQR) | 12.3 (7.93, 18.7) | 0.98 (0.7, 1.43) |  |
| Range | 1.92, 74.38 | 0.42, 79.92 |  |
| **TAT<sup>b</sup> (hours) by PCR SARS-CoV-2 result<sup>d</sup>** |  |  |  |
| Positive, n (%) | 538 (12.40%) | 638 (14.05%) | **<0.0001** |
| Mean (SD) | 13.5 (7.45) | 1.18 (1.04) |  |
| Median (IQR) | 12.26 (7.87, 18.73) | 0.93 (0.72, 1.33) |  |
| Range | 2.08, 74.38 | 0.43, 17.07 |  |
| Negative, n (%) | 3789 (87.32%) | 3902 (85.95%) | **<0.0001** |
| Mean (SD) | 13.27 (6.66) | 1.31 (1.88) |  |
| Median (IQR) | 12.27 (7.95, 18.65) | 0.98 (0.7, 1.45) |  |
| Range | 1.92, 61.7 | 0.42, 79.92 |  |
| Missing | 0 (0.00%) | 0 (0.00%) |  |
| Invalid/other, n (%) | 12 (0.28%) | 0 (0.00%) | **N/A** |
| Mean (SD) | 21.01 (5.82) | N/A |  |
| Median (IQR) | 19.73 (15.83, 26.575) | N/A |  |
| Range | 14.2, 28.73 | N/A |  |

This table may include multiple visits per patient. All individuals with symptoms of a possible SARS-CoV-2 infection, and a PCR SARS-CoV-2 test ordered by a clinician during the control period (April 2020–October 2020) or the intervention period (December 2020– May 2021), were considered eligible for this study and are included in this summary table.
<sup>a</sup>Wilcoxon rank-sum test
<sup>b</sup>TAT was calculated as the difference between test order date–time and result date–time
<sup>c</sup>Test excludes 'Invalid/other' category
<sup>d</sup>PCR SARS-CoV-2 test was the cobas SARS-CoV-2 test on the 6800 System or ePlex SARS-CoV-2
Test during the control period, and was the Liat SARS-CoV-2 & Influenza A/B test during the
intervention period
IQR, interquartile range; N/A, not applicable; PCR, polymerase chain reaction; POC, point-of-care;
SARS-CoV-2, severe acute respiratory syndrome coronavirus 2; SD, standard deviation

### Impact of rapid POC testing on ED LOS

Median ED LOS was longer in the intervention period (7.56 vs 7.15 hours; p=0.02) and for patients positive for SARS-CoV-2 (p<0.0001) (**Table 3**). ED LOS was similar in both periods for patients negative for SARS-CoV-2. By ED disposition, median LOS was longer in the intervention period for those who were discharged after ED care (5.10 hours vs 4.43 hours; p<0.0001) but shorter for those admitted (13.50 hours vs 18.48 hours; p<0.0001). ED LOS was similar in both periods for those who were admitted to the intensive care unit (ICU) or who died (both p>0.05) (**Table 3**).

**Table 3:** Summary of patient’s emergency department (ED) length of stay (LOS)

|  | **Control period** | **Intervention period** | **p-value<sup>a</sup>** |
| --- | --- | --- | --- |
|  | **(Centralized PCR)** | **(POC PCR)** |  |
| **Total patient visits, n** | **4339** | **4540** |  |
| **Patient's ED LOS (hours)** |  |  | 0.02<sup>b</sup> |
| Mean (SD) | 13.50 (16.49) | 13.30 (23.01) |  |
| Median (IQR) | 7.15 (4.08, 17.78) | 7.56 (4.62, 14.62) |  |
| Range | 0.32, 202.47 | 0.70, 776.88 |  |
| Missing | 0 (0.00%) | 0 (0.00%) |  |
| **ED LOS (hours) by PCR SARS-CoV-2 test result<sup>c</sup>** |  |  |  |
| Positive, n (%) | 538 (12.40%) | 638 (14.05%) | <0.0001 |
| Mean (SD) | 12.20 (14.79) | 21.57 (37.83) |  |
| Median (IQR) | 6.04 (3.10, 15.40) | 9.28 (4.60, 25.97) |  |
| Range | 0.53, 106.88 | 0.98, 383.27 |  |
| Missing | 0 (0.00%) | 0 (0.00%) |  |
| Negative, n (%) | 3789 (87.32%) | 3902 (85.95%) | 0.76 |
| Mean (SD) | 13.67 (16.69) | 11.95 (19.22) |  |
| Median (IQR) | 7.32 (4.20, 18.10) | 7.38 (4.63, 13.57) |  |
| Range | 0.32, 202.47 | 0.70, 776.88 |  |
| Missing | 0 (0.00%) | 0 (0.00%) |  |
| Invalid/other, n (%) | 12 (0.28%) | 0 (0.00%) | N/A |
| Mean (SD) | 17.37 (22.17) | N/A |  |
| Median (IQR) | 7.25 (2.85, 29.58) | N/A |  |
| Range | 1.15, 75.72 | N/A |  |
| Missing | 0 (0.00%) | 0 (0.00%) |  |
| **ED LOS (hours) by ED patient visit disposition<sup>c,d</sup>** |  |  |  |
| Discharge, n (%) | 2139 (49.30%) | 2258 (49.74%) | <0.0001 |
| Mean (SD) | 7.36 (11.69) | 8.20 (21.30) |  |
| Median (IQR) | 4.43 (2.67, 7.12) | 5.10 (3.35, 7.62) |  |
| Range | 0.32, 196.22 | 0.70, 776.88 |  |
| Hospital admission, n (%) | 1576 (36.32%) | 1740 (38.33%) | <0.0001 |
| Mean (SD) | 22.80 (19.07) | 20.10 (24.03) |  |
| Median (IQR) | 18.48 (8.74, 30.49) | 13.50 (8.43, 25.37) |  |
| Range | 1.53, 202.47 | 1.23, 383.27 |  |
| ICU admission, n (%) | 617 (14.22%) | 535 (11.78%) | 0.19 |
| Mean (SD) | 11.09 (12.30) | 12.83 (21.00) |  |
| Median (IQR) | 7.45 (5.02, 12.33) | 7.87 (5.50, 13.38) |  |
| Range | 1.23, 178.15 | 1.38, 346.48 |  |
| Death, n (%) | 7 (0.16%) | 6 (0.13%) | 1 |
| Mean (SD) | 8.65 (6.15) | 7.11 (2.88) |  |
| Median (IQR) | 7.80 (3.92, 10.83) | 6.51 (4.62, 9.27) |  |
| Range | 3.57, 21.32 | 4.12, 11.65 |  |

This table may include multiple visits per patient. All individuals with symptoms of a possible SARS- CoV-2 infection, and a PCR SARS-CoV-2 test ordered by a clinician during the control period (April 2020–October 2020) or the intervention period (December 2020– May 2021), were considered eligible for this study and are included in this summary table.
<sup>a</sup>Wilcoxon rank-sum test
<sup>b</sup>Test excludes 'Invalid/other' category
<sup>c</sup>PCR SARS-CoV-2 test was the cobas SARS-CoV-2 test on the 6800 System or the ePlex SARS- CoV-2 Test during the control period, and was the Liat SARS-CoV-2 & Influenza A/B test during the intervention period
<sup>d</sup>There were no data in the control period and only one record in the intervention period for 'Other' disposition category. ED LOS summary statistics for 'Other' disposition category were therefore not calculated
ICU, intensive care unit; IQR, interquartile range; N/A, not applicable; PCR, polymerase chain reaction; POC, point-of-care; SARS-CoV-2, severe acute respiratory syndrome coronavirus 2; SD, standard deviation

By SARS-CoV-2 test result and ED disposition, median ED LOS was longer in the intervention period for those who were discharged, regardless of test result (both p<0.0001) (**Supplementary Table S5**). For those admitted, median ED LOS was longer in the intervention period for patients with a positive SARS-CoV-2 result (p=0.02) and shorter with a negative result (p<0.0001).

Hospital LOS was similar between study periods overall and when analyzed by SARS-CoV-2 result (all comparisons, p>0.05; **Supplementary Table S6**). ICU LOS was shorter in the intervention period compared with the control period overall (2.08 days vs 2.49 days; p=0.01) and for those with a negative SARS-CoV-2 result (p=0.01) (**Supplementary Table S7**). Patients with a positive SARS-CoV-2 result had a similar ICU LOS in both periods (p>0.05).

### Impact of rapid POC testing on ED disposition

Fewer patients were admitted to the ICU in the intervention period (11.78%) compared with the control period (14.22%; p=0.0026) (**Table 4**). Patients with a positive SARS-CoV-2 result in the intervention period were more likely to be admitted to the hospital (p=0.0002) and less likely to be discharged (p=0.0002). Conversely, patients who tested negative were less likely to be admitted to the ICU in the intervention period (p=0.0006).

**Table 4:** Summary of emergency department (ED) disposition

|  | **Control period** | **Intervention period** | **p-value<sup>a</sup>** |
| --- | --- | --- | --- |
|  | **(Centralized PCR)** | **(POC PCR)** |  |
| **Total patient visits, n** | **4339** | **4540** |  |
| **ED disposition, n (%)** |  |  | 0.01<sup>b</sup> |
| Discharge | 2139 (49.30%) | 2258 (49.74%) | 0.72 |
| Hospital admission | 1576 (36.32%) | 1741 (38.35%) | 0.1 |
| ICU admission | 617 (14.22%) | 535 (11.78%) | 0.0026 |
| Death | 7 (0.16%) | 6 (0.13%) | 0.72 |
| Other | 0 (0.00%) | 1 (0.02%) | N/A |
| **ED disposition by PCR SARS-CoV-2 test result<sup>c</sup>, n, (%)** |  |  |  |
| **Positive** | 538 (12.40%) | 638 (14.05%) | 0.0005 |
| Discharge | 302 (56.13%) | 285 (44.67%) | 0.0002 |
| Hospital admission | 236 (43.87%) | 352 (55.17%) | 0.0002 |
| ICU admission | 69 (12.83%) | 85 (13.32%) | 0.8 |
| Death | 0 (0.00%) | 1 (0.16%) | N/A |
| Other | 0 (0.00%) | 0 (0.00%) | - |
| **Negative** | 3789 (87.32%) | 3902 (85.95%) | 0.0029 |
| Discharge | 1830 (48.30%) | 1973 (50.56%) | 0.09 |
| Hospital admission | 1952 (51.52%) | 1923 (49.28%) | 0.54 |
| ICU admission | 547 (14.44%) | 450 (11.53%) | 0.0006 |
| Death | 7 (0.18%) | 5 (0.13%) | 0.54 |
| Other | 0 (0.00%) | 1 (0.03%) | N/A |
| **Inconclusive** | 12 (0.28%) | 0 (0.00%) | N/A |
| Discharge | 7 (58.33%) | 0 (0.00%) | N/A |
| Hospital admission | 5 (41.67%) | 0 (0.00%) | N/A |
| ICU admission | 1 (8.33%) | 0 (0.00%) | N/A |
| Death | 0 (0.00%) | 0 (0.00%) | - |
| Other | 0 (0.00%) | 0 (0.00%) | - |

This table may include multiple visits per patient. All individuals with symptoms of a possible SARS- CoV-2 infection, and a PCR SARS-CoV-2 test ordered by a clinician during the control period (April
2020–October 2020) or the intervention period (December 2020– May 2021), were considered eligible for this study and are included in this summary table.
<sup>a</sup>Chi-square test. Subgroup p-values were adjusted for multiplicity using the false discovery rate method
<sup>b</sup>Test excludes ‘Other’ disposition category
<sup>c</sup>PCR SARS-CoV-2 test was the cobas SARS-CoV-2 test on the 6800 System or the ePlex SARS- CoV-2 Test during the control period, and was the Liat SARS-CoV-2 & Influenza A/B test during the intervention period
ICU, intensive care unit; N/A, not applicable; PCR, polymerase chain reaction; POC, point-of-care; SARS-CoV-2, severe acute respiratory syndrome coronavirus 2

### Impact of rapid POC testing on antibiotic/antiviral use

Antibiotic-only use was 14.5% lower in the intervention period (38.11% vs 44.55%; p<0.0001; **Table 5**), and the magnitude of the change was greater in patients with a positive vs negative SARS-CoV-2 result (47.2% vs 10.6% reduction; p<0.0001 for both). Antiviral-only use was higher in the intervention period compared with the control period (3.13% vs 0.94%; p<0.0001). Antivirals were prescribed more in the intervention period when patients were SARS-CoV-2 positive (14.89%) than when negative (0.92%). Approximately half of patients received no antibiotic or antiviral in both periods, though a higher proportion of patients received neither in the intervention period (54.25% vs 49.97%; p<0.0001).

**Table 5:** Summary of anti-infective prescriptions

|  | **Control period** | **Intervention period** | **p-value<sup>a</sup>** |
| --- | --- | --- | --- |
|  | **(Centralized PCR)** | **(POC PCR)** |  |
| **Total patient visits, n** | 4339 | 4540 |  |
| **Anti-infective prescription n, (%)** |  |  | <0.0001 |
| Antibiotic only | 1933 (44.55%) | 1730 (38.11%) | <0.0001 |
| Antiviral only | 38 (0.94%) | 131 (3.13%) | <0.0001 |
| Antibiotic and antiviral | 200 (4.61%) | 216 (4.76%) | 0.74 |
| No antibiotic or antiviral | 2168 (49.97%) | 2463 (54.25%) | <0.0001 |
| **Anti-infective prescription by test result<sup>b</sup>, n, (%)** |  |  |  |
| **Positive** | 538 (12.40%) | 638 (14.05%) | <0.0001 |
| Antibiotic only | 158 (29.37%) | 99 (15.52%) | <0.0001 |
| Antiviral only | 20 (3.72%) | 95 (14.89%) | <0.0001 |
| Antibiotic and antiviral | 83 (15.43%) | 129 (20.22%) | 0.04 |
| No antibiotic or antiviral | 277 (51.49%) | 315 (49.37%) | 0.47 |
| **Negative** | 3789 (87.32%) | 3902 (85.95%) | <0.0001 |
| Antibiotic only | 1771 (46.74%) | 1631 (41.80%) | <0.0001 |
| Antiviral only | 18 (0.48%) | 36 (0.92%) | 0.02 |
| Antibiotic and antiviral | 117 (3.09%) | 87 (2.23%) | 0.02 |
| No antibiotic or antiviral | 1883 (49.70%) | 2148 (55.05%) | <0.0001 |
| **Invalid/other** | 12 (0.28%) | 0 (0.00%) | N/A |
| Antibiotic only | 4 (33.33%) | 0 (0.00%) | N/A |
| Antiviral only | 0 (0.00%) | 0 (0.00%) | - |
| Antibiotic and antiviral | 0 (0.00%) | 0 (0.00%) | - |
| No antibiotic or antiviral | 8 (66.67%) | 0 (%) | N/A |
| **Anti-infective prescription by ED disposition<sup>c,d</sup>, n, (%)** |  |  |  |
| **Discharge** | 2139 (49.30%) | 2258 (49.74%) | 0.0004 |
| Antibiotic only | 437 (20.43%) | 388 (17.18%) | 0.02 |
| Antiviral only | 6 (0.28%) | 19 (0.84%) | 0.02 |
| Antibiotic and antiviral | 7 (0.33%) | 1 (0.04%) | 0.03 |
| No antibiotic or antiviral | 1689 (78.96%) | 1850 (81.93%) | 0.02 |
| **Hospital admission** | 1576 (36.32%) | 1740 (38.33%) | <0.0001 |
| Antibiotic only | 1025 (65.04%) | 970 (55.75%) | <0.0001 |
| Antiviral only | 27 (1.71%) | 101 (5.80%) | <0.0001 |
| Antibiotic and antiviral | 126 (7.99%) | 155 (8.91%) | 0.35 |
| No antibiotic or antiviral | 398 (25.25%) | 514 (29.54%) | 0.01 |
| **ICU admission** | 617 (14.22%) | 535 (11.78%) | 0.02 |
| Antibiotic only | 467 (75.69%) | 369 (68.97%) | 0.03 |
| Antiviral only | 5 (0.81%) | 11 (2.06%) | 0.1 |
| Antibiotic and antiviral | 67 (10.86%) | 60 (11.21%) | 0.85 |
| No antibiotic or antiviral | 78 (12.64%) | 95 (17.76%) | 0.03 |
| **Death** | 7 (0.16%) | 6 (0.13%) | 0.8 |
| Antibiotic only | 4 (57.14%) | 3 (50.00%) | 0.8 |
| Antiviral only | 0 (0.00%) | 0 (0.00%) | - |
| Antibiotic and antiviral | 0 (0.00%) | 0 (0.00%) | - |
| No antibiotic or antiviral | 3 (42.86%) | 3 (50.00%) | 0.8 |

One patient may receive multiple anti-infective prescriptions per visit. Antibiotic or antiviral prescriptions are counted as one instance per visit. All individuals with symptoms of a possible SARS-CoV-2 infection, and a PCR SARS-CoV-2 test ordered by a clinician during the control period (April 2020–October 2020) or the intervention period (December 2020– May 2021), were considered eligible for this study and are included in this summary table.
<sup>a</sup>Chi-square test. Subgroup p-values were adjusted for multiplicity using the false discovery rate method
<sup>b</sup>PCR SARS-CoV-2 test was the cobas SARS-CoV-2 test on the 6800 System or the ePlex SARS-CoV-2 Test during the control period, and was the Liat SARS-CoV-2 & Influenza A/B test during the intervention period
<sup>c</sup>There were no data in the control period and only one record in the intervention period for 'Other' disposition category. ED LOS summary statistics for 'Other' disposition category were therefore not calculated
ED, emergency department; ICU, intensive care unit; N/A, not applicable; PCR, polymerase chain reaction; POC, point-of-care; SARS-CoV-2, severe acute respiratory syndrome coronavirus 2

Patterns of anti-infective prescription were similar when analyzed by ED disposition (**Table 5**). However, as would be expected based on prescribing requirements, greater prescription of antivirals in the intervention period compared with the control period was more evident in those admitted to hospital or the ICU. By SARS-CoV-2 test result and ED disposition (**Supplementary Table S8**), antibiotic-only use in the intervention period compared with the control period was considerably lower in positive patients who were discharged or admitted (both p<0.0001). For SARS-CoV-2-negative patients, antibiotic prescriptions were similar in both periods for those who were discharged (p>0.05) but lower in the intervention period for those who were admitted to hospital (p=0.01).

Summaries of antibiotic and antiviral prescription rates by month are available in **Supplementary Tables S9 and S10**, respectively.

## Discussion

In this large study, ED patients with signs or symptoms consistent with COVID-19 received POC PCR test results in under an hour, compared with approximately 12 hours for centralized tests, when averaged across the cobas SARS-CoV-2 test and ePlex Test. The faster TAT did not reduce median ED LOS overall but did appear to reduce ED LOS for patients with negative test results who were subsequently admitted, suggesting more effective patient triage. The rate of antibiotic prescribing was also lower in the intervention period than in the control period overall, and in the population of patients with a positive SARS-CoV-2 result, which could support improved antibiotic stewardship with faster diagnosis.

Previous studies have evaluated the clinical performance of the cobas Liat SARS-CoV-2 & Influenza A/B test compared with other RT-PCR systems, such as the Cepheid^®^ GeneXpert system^®^ (26) and the cobas SARS-CoV-2 test on the cobas 6800/8800 Systems (27), and have demonstrated 100% positive percent agreement and 97–100% negative percent agreement. One study, conducted in an ED in Germany, estimated that POC PCR results using the cobas Liat System were available after 102 minutes from admission, which was shorter than the 811 minutes with central laboratory PCR (28). Though the TAT observed in the clinic is longer than the 20-minute assay run time, demonstrating some delay in the testing strategy, time to result is still substantially shorter than for centralized PCR methods.

Fast TAT with a reliable POC PCR assay for respiratory pathogens can reduce ED LOS (29). However, as in our study, some have reported increased ED LOS following the introduction of rapid diagnostic tests (30, 31). It is likely that factors other than TAT, such as availability of hospital resources (including number of isolation rooms), severity of symptoms, and presence of comorbidities requiring further work-up, contribute to longer ED LOS (30, 31). In this study, the ED had a higher number of symptomatic patient visits and positive SARS-CoV-2 test results in the intervention period than in the control period, likely related to the emergence of several SARS-CoV-2 variants, most notably Delta, diving surges (32), which may have contributed to increased LOS in the intervention period. Despite this, we found that ED LOS for patients with negative test results who were admitted was shorter following POC PCR implementation, indicating more effective patient triage.

Across both study periods, approximately 50% of ED patients were discharged and 37% were admitted. Though ICU admissions were lower in the intervention period, this result should be interpreted with care. Lower ICU admissions could indicate improved patient triage and management, which would have cost-saving implications (33, 34), but it is possible that patients in the control period had a variant associated with more severe disease (35), or that the reduction was due to other changes in hospital practices. Admission rates appeared higher than in other studies of symptomatic patients who were PCR tested for SARS-CoV-2, where admission rates were approximately 27–28% (36, 37). Higher admission rates could reflect the severity of symptoms encountered in this cohort, differences in hospital practices, or factors such as comorbidities that have not been controlled for. The UC Davis Medical Center is also a quaternary referral center, and the ED serves as a safety net for vulnerable populations and those with comorbidities of concern.

Several studies have demonstrated that implementing rapid POC PCR tests can improve clinical decision-making and management of patients with respiratory infections (27, 38, 39). Shorter time to diagnosis can reduce inappropriate antibiotic prescribing and allow for appropriate de-escalation (29, 38-41). Within the first 6 months of the pandemic, antibiotic prescribing rates for patients with COVID-19 was high (almost 75%), even in the absence of bacterial co-infection (42). In our study, though antibiotics were frequently prescribed, the rate was lower in the intervention period than in the control period, particularly in those with a positive SARS-CoV-2 result, supporting the notion that reduced time to result can improve antibiotic stewardship and help slow the emergence of antibiotic resistance (43, 44). Antiviral use was higher in the intervention period, though prescribing rates were low (<5%), likely reflecting the availability of antivirals and the stringent prescribing requirements at the time of the study.

One of the major limitations of this study is that due to the real-word, pre-post study design, several epidemiological factors could not be controlled for. The control and intervention periods were very different in terms of the stage of the COVID-19 pandemic. The control period coincided largely with the first wave, which peaked at approximately 10,000 new cases/day in mid-July (45). The intervention period was initiated in November, which was the start of a surge in cases in California, with approximately 47,000 new cases/day at the end of December and >100,000 cases/day in January (45). The study may also have been impacted by seasonality, with higher rates of respiratory infections generally encountered in winter (the intervention period) (46). In addition to environmental factors, COVID-19 treatment guidelines or practices were rapidly evolving (47, 48), and the availability of effective treatments changed over time (49), making clinical comparisons challenging.

Though the demographics of our patient population were not statistically different between periods, other non-collected characteristics of patients presenting to the ED may have differed (50), particularly as ED volumes were low during the beginning of the pandemic.

Nevertheless, this study provides an important insight into the outcomes observed following the implementation of a rapid POC SARS-CoV-2 test at a time when new cases of COVID-19 were high. It is also important to note that while our study supports the use of POC PCR tests in an emergency care setting, high-throughput centralized testing for SARS-CoV-2 remains an important diagnostic tool for patients not in need of immediate results. In this study, system identifiers were missing from the dataset, so it was not possible to analyze TAT for the cobas 6800 or ePlex systems separately; however, it is recognized that TAT using the ePlex SARS-CoV-2 Test is shorter than the cobas SARS-CoV-2 test. The ability to perform tests on the cobas Liat System, or other POC platforms, may increase capacity for testing to be conducted on centralized systems with broader respiratory pathogen testing panels, potentially improving testing efficiency and antibiotic prescribing practices.

## Conclusion

In conclusion, this study confirms that the cobas Liat SARS-CoV-2 & Influenza A/B test provides a considerably shorter TAT compared with centralized laboratory PCR methods, particularly those using sample batching, in a real-world emergency care setting. Decreased TAT also has the potential to reduce ED LOS in some patient populations and allows for better patient management and informed treatment decisions, in-line with improved antibiotic stewardship.

## Acknowledgements

The authors would like to thank Christopher Dodoo of Roche Molecular Systems, Inc., Pleasanton, California, USA, who contributed statistical support; Jeffrey Flack of University of California, Davis, California, USA for support with dataset creation; and Babar Javed of Roche Molecular Systems, Inc., Pleasanton, California, USA, who was the study manager. Medical writing support was provided by Samantha Forster of Elements Communications Ltd (Westerham, UK) and was funded by Roche Molecular Systems, Inc.

In the US, the cobas SARS-CoV-2 & Influenza A/B test (17) is only for use under the Food and Drug Administration’s Emergency Use Authorization. COBAS, LIAT, EPLEX and GENMARK DX are trademarks of Roche. All other product names and trademarks are the property of their respective owners.

---

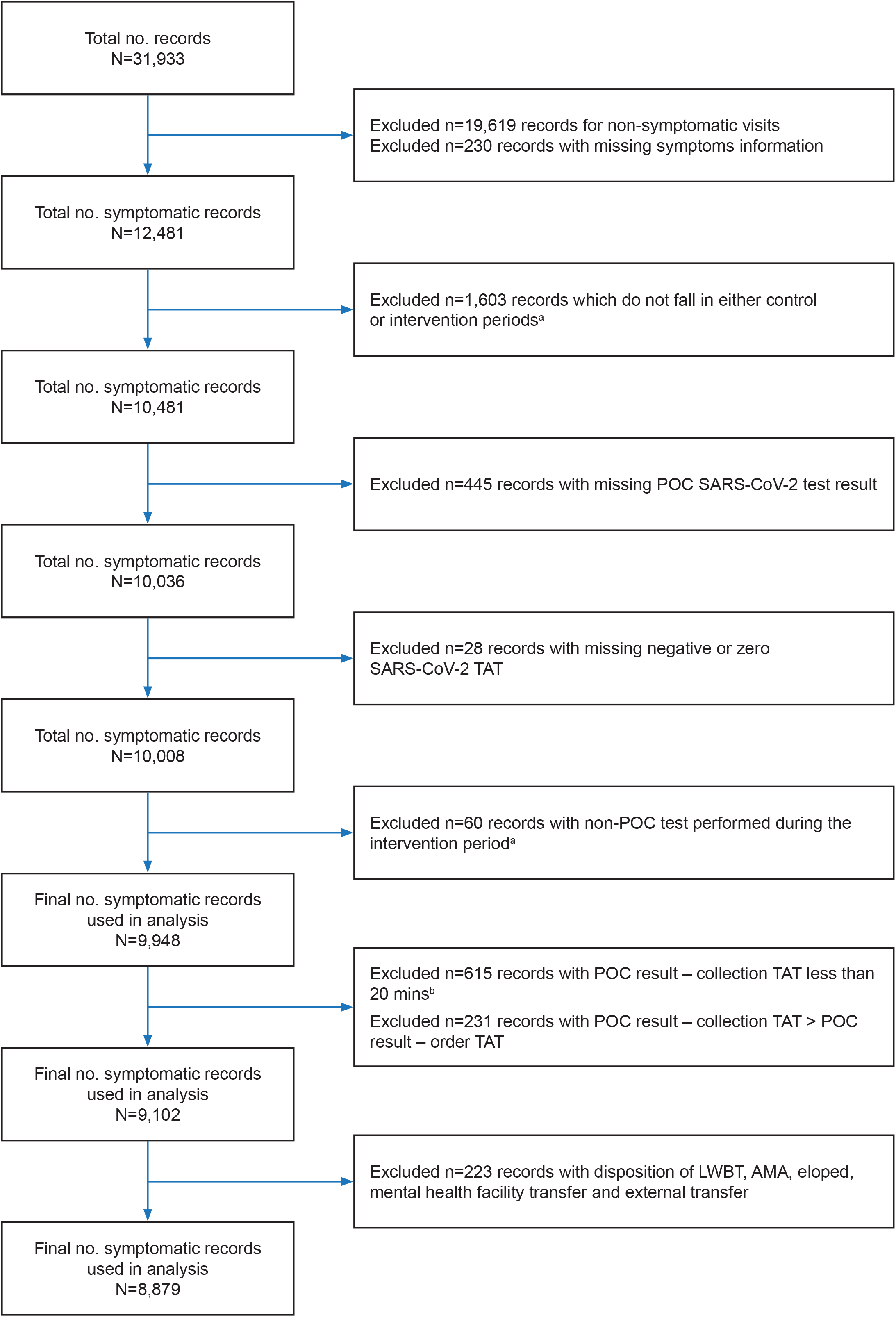

