## Supplementary Materials for "A study to assess the impact of cobas Liat point-of-care PCR assays (SARS-CoV-2 and Influenza A/B) on patient clinical management in the emergency department of the University of California at Davis Medical Center"

### Supplementary Information

Test methods

#### cobas® SARS-CoV-2 Test on the cobas 6800 System (control period; centralized testing)

The cobas SARS-CoV-2 Test for use on the cobas 6800/8800 Systems is an RT-PCR assay intended for the nucleic acids from SARS-CoV-2 in clinician-instructed, self-collected nasal swab specimens and clinician-collected nasal nasopharyngeal swab (NPS) and oropharyngeal swab samples from patients with signs and symptoms suggestive of COVID-19 (19). Samples were processed and analyzed as per the manufacturer’s instructions (19).

#### GenMark Dx® ePlex® SARS-CoV-2 Test (control period; centralized testing)

The GenMark Dx ePlex SARS-CoV-2 Test is an automated, RT-PCR assay intended for the qualitative detection of SARS-CoV-2 nucleic acid in NPS specimens (18). Specimens were collected from individuals suspected of COVID-19 by their healthcare provider and processed analyzed as per the manufacturer’s instructions for use (18).

#### cobas SARS-CoV-2 & Influenza A/B nucleic acid test for use on the cobas Liat® System (intervention period; POC PCR)

The cobas SARS-CoV-2 & Influenza A/B nucleic acid test for use on the cobas Liat System (Liat SARS-CoV-2 & Influenza A/B test) is an automated multiplex RT-PCR assay that integrates sample purification, nucleic acid amplification, and detection of the target sequences (17). In brief, the test utilizes provider-collected NPS and nasal swab specimens, and self-collected nasal swabs (collected in a healthcare setting with instruction by a healthcare provider) from individuals suspected of respiratory viral infection consistent with COVID-19. Samples were processed analyzed as per the instructions for use (17).

Analysis

Samples yielding indeterminate/invalid results from the cobas Liat SARS-CoV-2 & Influenza A/B test were re-run on the cobas Liat System, according to the IFU (17). Samples yielding indeterminate/invalid results from the cobas or GenMark Dx ePlex SARS-CoV-2 Tests were retested on the respective systems per their IFUs (18, 19). Following a second indeterminate/invalid result, cobas Liat and ePlex samples were run on the cobas 6800 System.

Tables

Supplementary Table S1: SARS-CoV-2-related symptoms

| | **Control period (Centralized PCR)** | **Intervention period (POC PCR)** |
|---|---|---|
| **Total patient visits, n** | **4339** | **4540** |
| **SARS-CoV-2-related symptoms, n (%)** | | |
| Shortness of breath/difficulty breathing | 947 (21.83%) | 843 (18.57%) |
| Fever | 621 (14.31%) | 453 (9.98%) |
| Cough | 338 (7.79%) | 266 (5.86%) |
| Abdominal pain | 287 (6.61%) | 352 (7.75%) |
| Vomiting | 199 (4.59%) | 224 (4.93%) |
| Headache | 118 (2.72%) | 112 (2.47%) |
| Sore throat | 111 (2.56%) | 89 (1.96%) |
| Nausea | 104 (2.40%) | 97 (2.14%) |
| Flu symptoms | 96 (2.21%) | 51 (1.12%) |
| Diarrhea | 85 (1.96%) | 61 (1.34%) |
| Chills | 44 (1.01%) | 21 (0.46%) |
| Myalgia (muscle or body aches) | 30 (0.69%) | 29 (0.64%) |
| Other symptoms | 21 (0.48%) | 18 (0.40%) |
| Loss of taste or smell | 10 (0.23%) | 9 (0.20%) |
| Symptoms not specified | 1760 (40.56%) | 2191 (48.26%) |
| Total[a] | | |

Percentages are based on the total patient visits. All individuals with symptoms of a possible SARS-CoV-2 infection, and a PCR SARS-CoV-2 test ordered by a clinician during the control period (April 2020–October 2020) or the intervention period (December 2020– May 2021), were considered eligible for this study and are included in this summary table.

[a]Includes ‘Symptoms not specified’

PCR, polymerase chain reaction; POC, point-of-care; SARS-CoV-2, severe acute respiratory syndrome coronavirus 2

Supplementary Table S2: Number of symptoms

| | Control period (Centralized PCR) | Intervention period (POC PCR) |
|---|---|---|
| **Total patient visits, n** | **4339** | **4540** |
| **SARS-CoV-2-related symptoms, n (%)** | | |
| 1 | 2365 (54.51%) | 2178 (47.97%) |
| 2 | 105 (2.42%) | 101 (2.22%) |
| 3 or more symptoms | 109 (2.51%) | 70 (1.54%) |
| Symptoms not specified | 1760 (40.56%) | 2191 (48.26%) |

Percentages are based on the total patient visits. All individuals with symptoms of a possible SARS-CoV-2 infection, and a PCR SARS-CoV-2 test ordered by a clinician during the control period (April 2020–October 2020) or the intervention period (December 2020– May 2021), were considered eligible for this study and are included in this summary table.

PCR, polymerase chain reaction; POC, point-of-care; SARS-CoV-2, severe acute respiratory syndrome coronavirus 2

Supplementary Table S3: PCR SARS-CoV-2 test results

| | Control period<br><br>(Centralized PCR) | Intervention period<br><br>(POC PCR) | p-value[a] |
|---|---|---|---|
| **Total patient visits, n** | **4339** | **4540** | |
| **PCR SARS-CoV-2 test,[b] n (%)** | | | |
| Positive | 538 (12.40%) | 638 (14.05%) | 0.02[c] |
| Negative | 3789 (87.32%) | 3902 (85.95%) | |
| Invalid/other | 12 (0.28%) | 0 (0.00%) | |

Percentages are based on the total patient visits. All individuals with symptoms of a possible SARS-CoV-2 infection, and a PCR SARS-CoV-2 test ordered by a clinician during the control period (April 2020–October 2020) or the intervention period (December 2020– May 2021), were considered eligible for this study and are included in this summary table.

[a]Chi-square test

[b]PCR SARS-CoV-2 test was the cobas SARS-CoV-2 test on the 6800 System or the ePlex SARS-CoV-2 Test during the control period, and was the Liat SARS-CoV-2 & Influenza A/B test during the intervention period. Only visits with non-missing result values are included in this summary table

[c]Test excludes ‘Invalid/other’ category

PCR, polymerase chain reaction; POC, point-of-care; SARS-CoV-2, severe acute respiratory syndrome coronavirus 2

Supplementary Table S4: Impact on turnaround time (TAT), collection to result

| | **Control period**<br><br>**(Centralized PCR)** | **Intervention period**<br><br>**(POC PCR)** | **p-value[a]** |
|---|---|---|---|
| **Total patient visits, n** | 4339 | 4540 | |
| **TAT[b] (hours)** | | | <0.0001[c] |
| Mean (SD) | 13.17 (6.77) | 1.16 (1.79) | |
| Median (IQR) | 12.15 (7.8, 18.55) | 0.83 (0.57, 1.3) | |
| Range | 1.83, 74.23 | 0.33, 79.75 | |
| **TAT[b] (hours) by PCR SARS-CoV-2 result[d]** | | | |
| Positive, n (%) | 538 (12.40%) | 638 (14.05%) | <0.0001 |
| Mean (SD) | 13.34 (7.45) | 1.05 (1.04) | |
| Median (IQR) | 12.08 (7.72, 18.57) | 0.8 (0.58, 1.22) | |
| Range | 1.95, 74.23 | 0.33, 16.9 | |
| Negative, n (%) | 3789 (87.32%) | 3902 (85.95%) | <0.0001 |
| Mean (SD) | 13.12 (6.65) | 1.17 (1.88) | |
| Median (IQR) | 12.12 (7.82, 18.5) | 0.85 (0.57, 1.32) | |
| Range | 1.83, 61.6 | 0.33, 79.75 | |
| Invalid/other, n (%) | 12 (0.28%) | 0 (0.00%) | N/A |
| Mean (SD) | 20.88 (5.84) | N/A | |
| Median (IQR) | 19.61 (15.695, 26.46) | N/A | |
| Range | 14.03, 28.6 | N/A | |

This table may include multiple visits per patient. All individuals with symptoms of a possible SARS-CoV-2 infection, and a PCR SARS-CoV-2 test ordered by a clinician during the control period (April 2020–October 2020) or the intervention period (December 2020– May 2021), were considered eligible for this study and are included in this summary table.

[a]Wilcoxon rank-sum test

[b]TAT was calculated as difference between sample collection date–time and result date–time

[c]Test excludes 'Invalid/other' category

[d]PCR SARS-CoV-2 test was the cobas SARS-CoV-2 test on the 6800 System or the ePlex SARS-CoV-2 Test during the control period, and was the Liat SARS-CoV-2 & Influenza A/B test during the intervention period

IQR, interquartile range; N/A, not applicable; PCR, polymerase chain reaction; POC, point-of-care; SARS-CoV-2, severe acute respiratory syndrome coronavirus 2; SD, standard deviation

Supplementary Table S5: Summary of patient's emergency department (ED) length of stay (LOS) by SARS-CoV-2 test result and ED disposition

| | Control period (Centralized PCR) | Intervention period (POC PCR) | p-value[a] |
|---|---|---|---|
| **Total patient visits, n** | **4339** | **4540** | |
| **Patient's ED LOS (hours)** | | | 0.02[b] |
| Mean (SD) | 13.50 (16.49) | 13.30 (23.01) | |
| Median (IQR) | 7.15 (4.08, 17.78) | 7.56 (4.62, 14.62) | |
| Range | 0.32, 202.47 | 0.70, 776.88 | |
| Missing | 0 (0.00%) | 0 (0.00%) | |
| **ED LOS (hours) by ED patient visit disposition[c] and PCR SARS-CoV-2 test result[d,e]** | | | |
| **Positive**, n (%) | 538 (12.43%) | 638 (14.05%) | |
| Discharge, n (%) | 302 (56.13%) | 285 (44.67%) | <0.0001 |
| Mean (SD) | 4.99 (5.73) | 7.70 (18.08) | |
| Median (IQR) | 3.69 (2.02, 5.72) | 4.53 (2.92, 7.42) | |
| Range | 0.53, 46.37 | 0.98, 282.95 | |
| Hospital admission, n (%) | 167 (31.04%) | 267 (41.85%) | 0.02 |
| Mean (SD) | 24.63 (18.74) | 35.56 (44.73) | |
| Median (IQR) | 21.62 (9.42, 31.20) | 25.40 (11.40, 40.22) | |
| Range | 2.20, 106.88 | 3.48, 383.27 | |
| ICU admission, n (%) | 69 (12.83%) | 85 (13.32%) | 0.39 |
| Mean (SD) | 13.73 (10.57) | 24.32 (46.11) | |
| Median (IQR) | 11.02 (5.48, 17.78) | 10.07 (6.17, 26.10) | |
| Range | 2.60, 50.62 | 1.88, 346.48 | |
| Death, n (%) | 0 (0%) | 1 (0.16%) | N/A |
| Mean (SD) | N/A | N/A | |
| Median (IQR) | N/A | N/A | |
| Range | N/A | N/A | |
| **Negative**, n (%) | 3789 (87.57%) | 3902 (85.95%) | |
| Discharge, n (%) | 1830 (48.3%) | 1973 (50.56%) | <0.0001 |
| Mean (SD) | 7.76 (12.38) | 8.27 (21.73) | |
| Median (IQR) | 4.59 (2.85, 7.43) | 5.18 (3.40, 7.65) | |

| | | | |
|---|---|---|---|
| Range | 0.32, 196.22 | 0.70, 776.88 | |
| Hospital admission, n (%) | 1405 (37.08%) | 1473 (37.75%) | <0.0001 |
| Mean (SD) | 22.52 (19.08) | 17.30 (16.42) | |
| Median (IQR) | 17.93 (8.65, 30.35) | 12.53 (8.12, 23.47) | |
| Range | 1.53, 202.47 | 1.23, 371.28 | |
| ICU admission, n (%) | 547 (14.44%) | 450 (11.53%) | 0.4 |
| Mean (SD) | 10.76 (12.48) | 10.66 (9.85) | |
| Median (IQR) | 7.22 (4.97, 11.80) | 7.59 (5.35, 12.57) | |
| Range | 1.23, 178.15 | 1.38, 92.90 | |
| Death, n (%) | 7 (0.18%) | 5 (0.13%) | 0.87 |
| Mean (SD) | 8.65 (6.15) | 7.71 (2.77) | |
| Median (IQR) | 7.80 (3.92, 10.83) | 6.90 (6.12, 9.27) | |
| Range | 3.57, 21.32 | 4.62, 11.65 | |

This table may include multiple visits per patient. All individuals with symptoms of a possible SARS-CoV-2 infection, and a PCR SARS-CoV-2 test ordered by a clinician during the control period (April 2020–October 2020) or the intervention period (December 2020– May 2021), were considered eligible for this study and are included in this summary table.

[a]Wilcoxon rank-sum test

[b]Test excludes ‘Invalid/other’ category

[c]There were no data in the control period and only one record in the intervention period for ‘Other’ disposition category. ED LOS summary statistics for 'Other' disposition category were therefore not calculated

[d]PCR SARS-CoV-2 test was the cobas SARS-CoV-2 test on the 6800 System or the ePlex SARS-CoV-2 Test during the control period, and was the Liat SARS-CoV-2 & Influenza A/B test during the intervention period

[e]Data from invalid test results are not included in this table. There were only 12 invalid test results from the control period and none from the intervention period

ICU, intensive care unit; IQR, interquartile range; N/A, not applicable; PCR, polymerase chain reaction; POC, point-of-care; SARS-CoV-2, severe acute respiratory syndrome coronavirus 2; SD, standard deviation

Supplementary Table S6: Summary of patient's hospital length of stay (LOS)

| | **Control period (Centralized PCR)** | **Intervention period (POC PCR)** | **p-value[a]** |
|---|---|---|---|
| **Total patient visits for patients admitted to hospital, n** | **1576** | **1737** | |
| **Patient's hospital LOS (days)** | | | 0.66[b] |
| Mean (SD) | 5.22 (11.66) | 4.85 (5.93) | |
| Median (IQR) | 3.06 (1.71, 5.83) | 3.09 (1.71, 5.86) | |
| Range | 0.02, 387.80 | 0.02, 116.92 | |
| **Hospital LOS (days) by PCR SARS-CoV-2 test result[c]** | | | |
| Positive, n (%) | 167 (10.60%) | 267 (15.34%) | 0.58 |
| Mean (SD) | 6.49 (5.73) | 6.22 (5.92) | |
| Median (IQR) | 4.92 (2.21, 9.02) | 4.78 (2.73, 7.76) | |
| Range | 0.07, 43.55 | 0.11, 47.15 | |
| Negative, n (%) | 1405 (89.15%) | 1470 (84.48%) | 0.24 |
| Mean (SD) | 5.08 (12.18) | 4.61 (5.90) | |
| Median (IQR) | 2.95 (1.68, 5.47) | 2.84 (1.64, 5.63) | |
| Range | 0.02, 387.80 | 0.02, 116.92 | |
| Invalid/other, n (%) | 4 (0.25%) | 0 (0.00%) | N/A |
| Mean (SD) | 1.98 (0.55) | N/A | |
| Median (IQR) | 1.85 (1.66, 2.31) | N/A | |
| Range | 1.48, 2.76 | N/A | |

This table may include multiple visits per patient. All individuals with symptoms of a possible SARS-CoV-2 infection, and a PCR SARS-CoV-2 test ordered by a clinician during the control period (April 2020–October 2020) or the intervention period (December 2020– May 2021), were considered eligible for this study and are included in this summary table.

[a]Wilcoxon rank-sum test

[b]Test excludes 'Invalid/other' category

[c]PCR SARS-CoV-2 test was the cobas SARS-CoV-2 test on the 6800 System or the ePlex SARS-CoV-2 Test during the control period, and was the Liat SARS-CoV-2 & Influenza A/B test during the intervention period

IQR, interquartile range; N/A, not applicable; PCR, polymerase chain reaction; POC, point-of-care; SARS-CoV-2, severe acute respiratory syndrome coronavirus 2; SD, standard deviation

Supplementary Table S7: Summary of patient's intensive care unit (ICU) length of stay (LOS)

| | **Control period (Centralized PCR)** | **Intervention period (POC PCR)** | **p-value[a]** |
|---|---|---|---|
| **Total patient visits for patients admitted to ICU, n** | 617 | 535 | |
| **Patient's ICU LOS (in days)** | | | 0.01[b] |
| Mean (SD) | 5.65 (8.73) | 5.12 (9.09) | |
| Median (IQR) | 2.49 (1.23, 6.01) | 2.08 (1.03, 4.77) | |
| Range | 0.13, 82.00 | 0.00, 108.18 | |
| **ICU LOS (days) by PCR SARS-CoV-2 test result[c]** | | | |
| Positive, n (%) | 69 (11.18%) | 85 (15.89%) | 0.12 |
| Mean (SD) | 11.91 (13.58) | 10.96 (13.68) | |
| Median (IQR) | 5.94 (3.14, 16.28) | 4.34 (1.87, 14.81) | |
| Range | 0.74, 62.37 | 0.31, 52.57 | |
| Negative, n (%) | 547 (88.65%) | 450 (84.11%) | 0.01 |
| Mean (SD) | 4.86 (7.57) | 4.01 (7.45) | |
| Median (IQR) | 2.21 (1.12, 5.32) | 1.88 (0.99, 3.88) | |
| Range | 0.13, 82.00 | 0.00, 108.18 | |
| Invalid/other, n (%) | 1 (0.16%) | 0 (0.00%) | N/A |
| Mean (SD) | 4.05 (N/A) | N/A | |
| Median (IQR) | 4.05 (4.05, 4.05) | N/A | |
| Range | 4.05, 4.05 | N/A | |

This table may include multiple visits per patient. All individuals with symptoms of a possible SARS-CoV-2 infection, and a PCR SARS-CoV-2 test ordered by a clinician during the control period (April 2020–October 2020) or the intervention period (December 2020– May 2021), were considered eligible for this study and are included in this summary table.

[a]Wilcoxon rank-sum test

[b]Test excludes 'Invalid/other' category

[c]PCR SARS-CoV-2 test was the cobas SARS-CoV-2 test on the 6800 System or the ePlex SARS-CoV-2 Test during the control period, and was the Liat SARS-CoV-2 & Influenza A/B test during the intervention period

IQR, interquartile range; N/A, not applicable; PCR, polymerase chain reaction; POC, point-of-care; SARS-CoV-2, severe acute respiratory syndrome coronavirus 2; SD, standard deviation

Supplementary Table S8: Summary of anti-infective prescription by SARS-CoV-2 test result and emergency department (ED) disposition

| | Control period (Centralized PCR) | Intervention period (POC PCR) | p-value[a] |
|---|---|---|---|
| **Total patient visits, n** | **4339** | **4540** | |
| **Anti-infective prescription, n (%)** | | | <0.0001 |
| Antibiotic only | 1933 (44.55%) | 1730 (38.11%) | <0.0001 |
| Antiviral only | 38 (0.94%) | 131 (3.13%) | <0.0001 |
| Antibiotic and antiviral | 200 (4.61%) | 216 (4.76%) | 0.74 |
| No antibiotic or antiviral | 2168 (49.97%) | 2463 (54.25%) | <0.0001 |
| **Anti-infective prescription by PCR SARS-CoV-2 test result[b] and ED disposition,[c,d] n (%)** | | | |
| **Positive** | 538 (12.43%) | 638 (14.05%) | |
| Discharge | 302 (56.13%) | 285 (44.67%) | <0.0001 |
| Antibiotic only | 61 (20.2%) | 25 (8.77%) | <0.0001 |
| Antiviral only | 0 (0.00%) | 0 (0.00%) | - |
| Antibiotic and antiviral | 0 (0.00%) | 0 (0.00%) | - |
| No antibiotic or antiviral | 241 (79.8%) | 260 (91.23%) | <0.0001 |
| Hospital admission | 167 (31.04%) | 267 (41.85%) | <0.0001 |
| Antibiotic only | 77 (46.11%) | 54 (20.22%) | <0.0001 |
| Antiviral only | 16 (9.58%) | 87 (32.58%) | <0.0001 |
| Antibiotic and antiviral | 41 (24.55%) | 82 (30.71%) | 0.22 |
| No antibiotic or antiviral | 33 (19.76%) | 44 (16.48%) | 0.38 |
| ICU admission | 69 (12.83%) | 85 (13.32%) | 0.29 |
| Antibiotic only | 20 (28.99%) | 20 (23.53%) | 0.49 |
| Antiviral only | 4 (5.8%) | 8 (9.41%) | 0.49 |
| Antibiotic and antiviral | 42 (60.87%) | 47 (55.29%) | 0.49 |
| No antibiotic or antiviral | 3 (4.35%) | 10 (11.76%) | 0.4 |
| Death | 0 (0.00%) | 1 (0.16%) | N/A |
| Antibiotic only | 0 (0.00%) | 0 (0.00%) | - |
| Antiviral only | 0 (0.00%) | 0 (0.00%) | - |
| Antibiotic and antiviral | 0 (0.00%) | 0 (0.00%) | - |
| No antibiotic or antiviral | 0 (0.00%) | 1 (100%) | N/A |

| | | | |
|---|---|---|---|
| **Negative** | 3789 (87.57%) | 3902 (85.95%) | |
| Discharge | 1830 (48.3%) | 1973 (50.56%) | 0.0037 |
| Antibiotic only | 376 (20.55%) | 363 (18.4%) | 0.13 |
| Antiviral only | 6 (0.33%) | 19 (0.96%) | 0.05[e] |
| Antibiotic and antiviral | 7 (0.38%) | 1 (0.05%) | 0.05[f] |
| No antibiotic or antiviral | 1441 (78.74%) | 1590 (80.59%) | 0.16 |
| Hospital admission | 1405 (37.08%) | 1473 (37.75%) | 0.0035 |
| Antibiotic only | 945 (67.26%) | 916 (62.19%) | 0.01 |
| Antiviral only | 11 (0.78%) | 14 (0.95%) | 0.63 |
| Antibiotic and antiviral | 85 (6.05%) | 73 (4.96%) | 0.26 |
| No antibiotic or antiviral | 364 (25.91%) | 470 (31.91%) | 0.0016 |
| ICU admission | 547 (14.44%) | 450 (11.53%) | 0.05 |
| Antibiotic only | 446 (81.54%) | 349 (77.56%) | 0.22 |
| Antiviral only | 1 (0.18%) | 3 (0.67%) | 0.23 |
| Antibiotic and antiviral | 25 (4.57%) | 13 (2.89%) | 0.22 |
| No antibiotic or antiviral | 75 (13.71%) | 85 (18.89%) | 0.11[g] |
| Death | 7 (0.18%) | 5 (0.13%) | 0.92 |
| Antibiotic only | 4 (57.14%) | 3 (60%) | 0.92 |
| Antiviral only | 0 (0.00%) | 0 (0.00%) | - |
| Antibiotic and antiviral | 0 (0.00%) | 0 (0.00%) | - |
| No antibiotic or antiviral | 3 (42.86%) | 2 (40%) | 0.92 |

One patient may receive multiple anti-infective prescriptions per visit. Antibiotic or antiviral prescriptions are counted as one instance per visit. All individuals with symptoms of a possible SARS-CoV-2 infection, and a PCR SARS-CoV-2 test ordered by a clinician during the control period (April 2020–October 2020) or the intervention period (December 2020– May 2021), were considered eligible for this study and are included in this summary table.

[a]Chi-square test. Subgroup p-values were adjusted for multiplicity using the false discovery rate method

[b]PCR SARS-CoV-2 test was the cobas SARS-CoV-2 test on the 6800 System or the ePlex SARS-CoV-2 Test during the control period, and was the Liat SARS-CoV-2 & Influenza A/B test during the intervention period

[c]There were no data in the control period and only one record in the intervention period for ‘Other’ disposition category. Data from the 'Other' disposition category were therefore not calculated

[d]Data from invalid test results are not included in this table. There were only 12 invalid test results from the control period and none from the intervention period

[e]Unadjusted p-value of 0.02

[f]Unadjusted p-value of 0.03

[g]Unadjusted p-value of 0.03

ICU, intensive care unit; LOS, length of stay; N/A, not applicable; PCR, polymerase chain reaction; POC, point-of-care; SARS-CoV-2, severe acute respiratory syndrome coronavirus 2

Supplementary Table 9: Summary of antibiotic prescriptions by month

| Control period (Centralized PCR) | | | | | Intervention period (POC PCR) | | | | |
|---|---|---|---|---|---|---|---|---|---|
| **Patient group** | **Overall** | **PCR+** | **PCR-** | **PCR invalid** | **Patient group** | **Overall** | **PCR+** | **PCR-** | **PCR invalid** |
| **Overall antibiotic prescriptions, n (%)** | 6347 (100.00%) | 599 (9.44%) | 5737 (90.39%) | 11 (0.17%) | **Overall antibiotic prescriptions, n (%)** | 5713 (100.00%) | 665 (11.64%) | 5048 (88.36%) | 0 (0.00%) |
| April 2020 | 863 (13.60%) | 53 (6.14%) | 810 (93.86%) | 0 (0.00%) | December 2020 | 1326 (23.21%) | 260 (19.61%) | 1066 (80.39%) | 0 (0.00%) |
| May 2020 | 687 (10.82%) | 9 (1.31%) | 678 (98.69%) | 0 (0.00%) | January 2021 | 1365 (23.89%) | 243 (17.80%) | 1122 (82.20%) | 0 (0.00%) |
| June 2020 | 962 (15.16%) | 95 (9.88%) | 867 (90.12%) | 0 (0.00%) | February 2021 | 984 (17.22%) | 86 (8.74%) | 898 (91.26%) | 0 (0.00%) |
| July 2020 | 1167 (18.39%) | 178 (15.25%) | 987 (84.58%) | 2 (0.17%) | March 2021 | 837 (14.65%) | 36 (4.30%) | 801 (95.70%) | 0 (0.00%) |
| August 2020 | 1097 (17.28%) | 174 (15.86%) | 923 (84.14%) | 0 (0.00%) | April 2021 | 627 (10.97%) | 24 (3.83%) | 603 (96.17%) | 0 (0.00%) |
| September 2020 | 768 (12.10%) | 51 (6.64%) | 712 (92.71%) | 5 (0.65%) | May 2021 | 574 (10.05%) | 16 (2.79%) | 558 (97.21%) | 0 (0.00%) |
| October 2020 | 803 (12.65%) | 39 (4.86%) | 760 (94.65%) | 4 (0.50%) | | | | | |

All individuals with symptoms of a possible SARS-CoV-2 infection, and a PCR SARS-CoV-2 test ordered by a clinician during the control period (April 2020–October 2020) or the intervention period (December 2020– May 2021), were considered eligible for this study and are included in this summary table.

PCR, polymerase chain reaction; POC, point-of-care; SARS-CoV-2, severe acute respiratory syndrome coronavirus 2

Supplementary Table 10: Summary of antiviral prescriptions by month

| Control period (Centralized PCR) | | | | | Intervention period (POC PCR) | | | | |
|---|---|---|---|---|---|---|---|---|---|
| **Patient group** | **Overall** | **PCR+** | **PCR-** | **PCR invalid** | **Patient group** | **Overall** | **PCR+** | **PCR-** | **PCR invalid** |
| **Overall antiviral prescriptions, n (%)** | 315 (100.00%) | 108 (34.29%) | 207 (65.71%) | 0 (0.00%) | **Overall antiviral prescriptions, n (%)** | 419 (100.00%) | 236 (56.32%) | 183 (43.68%) | 0 (0.00%) |
| April 2020 | 44 (13.97%) | 11 (25.00%) | 33 (75.00%) | 0 (0.00%) | December 2020 | 134 (31.98%) | 100 (74.63%) | 34 (25.37%) | 0 (0.00%) |
| May 2020 | 23 (7.30%) | 3 (13.04%) | 20 (86.96%) | 0 (0.00%) | January 2021 | 108 (25.78%) | 72 (66.67%) | 36 (33.33%) | 0 (0.00%) |
| June 2020 | 47 (14.92%) | 16 (34.04%) | 31 (65.96%) | 0 (0.00%) | February 2021 | 68 (16.23%) | 33 (48.53%) | 35 (51.47%) | 0 (0.00%) |
| July 2020 | 53 (16.83%) | 16 (30.19%) | 37 (69.81%) | 0 (0.00%) | March 2021 | 40 (9.55%) | 15 (37.50%) | 25 (62.50%) | 0 (0.00%) |
| August 2020 | 49 (15.56%) | 30 (61.22%) | 19 (38.78%) | 0 (0.00%) | April 2021 | 37 (8.83%) | 8 (21.62%) | 29 (78.38%) | 0 (0.00%) |
| September 2020 | 59 (18.73%) | 17 (28.81%) | 42 (71.19%) | 0 (0.00%) | May 2021 | 32 (7.64%) | 8 (25.00%) | 24 (75.00%) | 0 (0.00%) |
| October 2020 | 40 (12.70%) | 15 (37.50%) | 25 (62.50%) | 0 (0.00%) | | | | | |

All individuals with symptoms of a possible SARS-CoV-2 infection, and a PCR SARS-CoV-2 test ordered by a clinician during the control period (April 2020–October 2020) or the intervention period (December 2020– May 2021), were considered eligible for this study and are included in this summary table.

PCR, polymerase chain reaction; POC, point-of-care; SARS-CoV-2, severe acute respiratory syndrome coronavirus 2

Figures

Supplementary Figure 1: Study flowchart

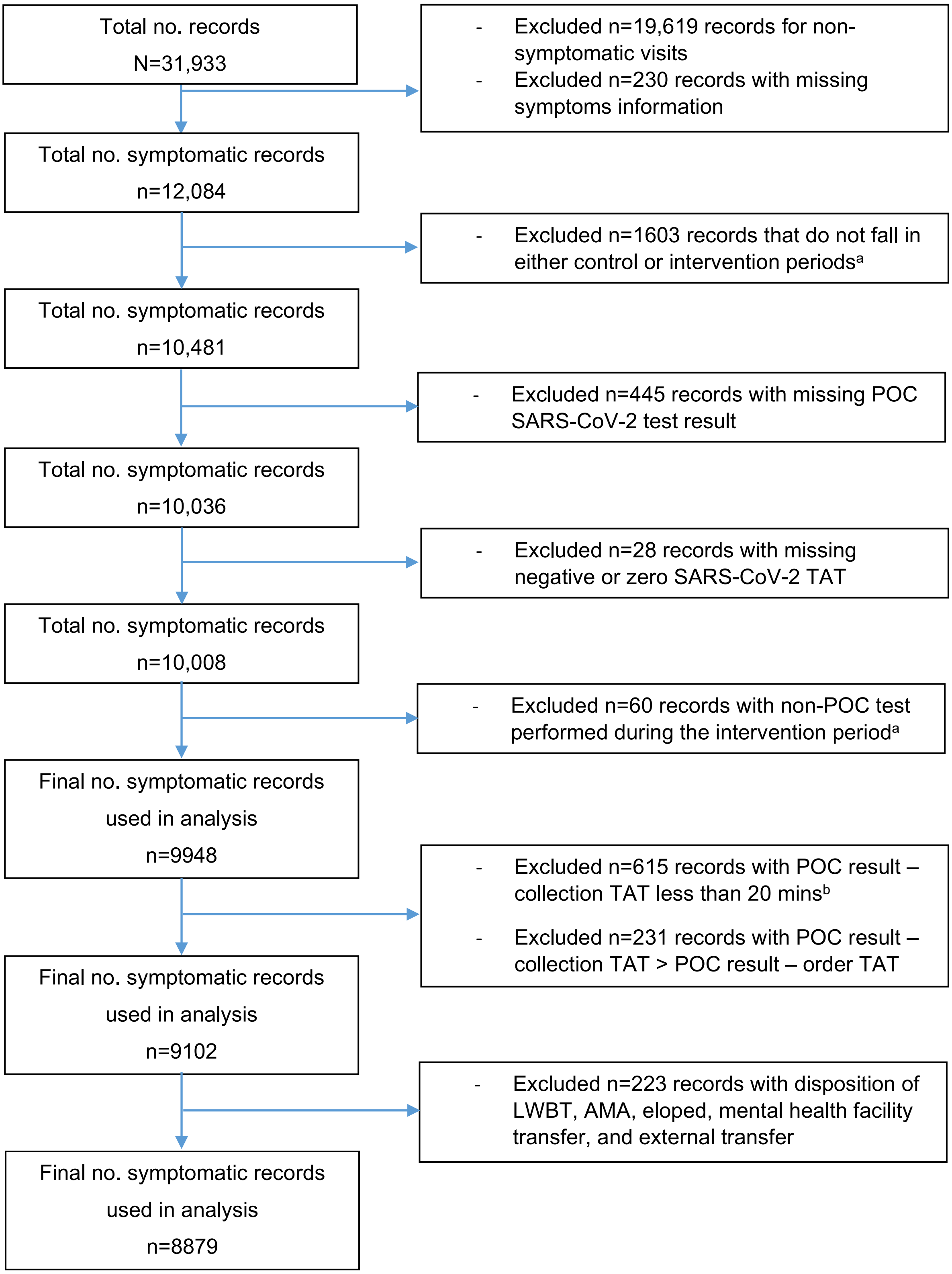

A total of 31,933 patient encounters were evaluated. Following exclusion based on predefined study criteria, 8879 (27.81%) medical records were included in the study.

[a]Control period: April 2020–October 2020; intervention period: December 2020– May 2021

[b]The Liat SARS-CoV-2 & Influenza A/B test takes a minimum of 20 minutes to complete

AMA, against medical advice; LWBT, left without being treated; POC, point-of-care; SARS-CoV-2, severe acute respiratory syndrome coronavirus 2; TAT, turnaround time
